# Dynamical Indicators of Resilience from Physiological Time Series in Geriatric Inpatients: Lessons Learned

**DOI:** 10.1101/2020.12.04.20243121

**Authors:** Jerrald L. Rector, Sanne M. W. Gijzel, Ingrid A. van de Leemput, Fokke B. van Meulen, Marcel G. M. Olde Rikkert, René J. F. Melis

## Abstract

**Background:** The concept of physical resilience may help geriatric medicine objectively assess patients’ ability to ‘bounce back’ from future health challenges. Indicators hypothesized to forecast resilience after a stressor have been developed under two paradigms with different perspectives: Critical Slowing Down (CSD) and Loss of Complexity (LoC). This study explored if and how these indicators, based on fluctuations in physiologic signals, can validly reflect the physical resilience of geriatric inpatients.

**Methods:** Geriatric patients (n = 121, 60% female) had their heart rate and physical activity continuously monitored using a chest-worn sensor. Measures of health functioning (multimorbidity, frailty and Activities of Daily Living [ADL]) were obtained by questionnaire at admission. Indicators from both paradigms (CSD: variance, autocorrelation, cross-correlation; LoC: [multivariate] multiscale entropy) were extracted from both physiological signals. The relationships among indicators and their associations with health functioning were assessed by correlation and linear regression analyses, respectively.

**Results:** Greater complexity and higher variance in physical activity were associated with lower frailty (β = –0.28, p=.004 and β = –0.37, p<.001, respectively) and better ADL function (β = 0.23, p=.022 and β = 0.38, p<.001). The associations of physical activity variance with health functioning was not in the expected direction based on the Critical Slowing Down paradigm.

**Conclusions:** Associations between dynamical resilience indicators tested here and measures of health functioning were not all in the expected direction. In retrospect, these observations stress the importance of matching the underlying assumptions of the resilience paradigm to the homeostatic role of the variable monitored.

## Introduction

The population is aging, and individuals in need of medical care are simultaneously becoming older, frailer and more likely to have multiple diseases (1). Even with detailed prognostic assessment and clinical intuitions, medicine is unable to objectively assess who will resist and recover from health stressors imposed by disease or by its treatment. The concept of physical resilience – the individual’s capacity to resist functional decline and recover physical health following a stressor – may facilitate this objective assessment and simultaneously shift medicine towards a more positive outlook emphasizing a person’s resources instead of deficits (2). In fact, the development of tools that inform clinical intuitions about older adults’ resilience has been termed a priority in aging research (3).

Researchers have developed a number of concepts and associated indicators that may help quantify the resilience of the older adult (4-8). The current study focuses on indicators hypothesized to be associated with resilient outcomes after a physical stressor. These indicators come from two leading paradigms that take modeling humans as a complex system as a starting point: the ‘Critical Slowing Down’ (CSD) and the ‘Loss of Complexity’ (LoC) paradigms.

The CSD approach proposes that as a generally stable system becomes less resilient, system variables show increasing delays in their recovery from internal or external perturbations (7,9). Specifically, with loss of resilience, physiological time series display larger fluctuations, slower recovery to equilibrium, and more rigid coupling between sub-systems (e.g., cardiovascular and respiratory systems). These indicators of CSD are reflected statistically in increased variance, temporal autocorrelation and cross-correlations between sub-systems, respectively (8). Empirical validation of the predictive validity of these CSD-related indicators in human psychology and physiology is emerging (10-13). By extracting these indicators from the patient’s dynamic responses to naturally occurring small-scale perturbations, or micro-perturbations, such as ingesting a meal or standing from a chair, one can make predictions about the response of the older adult to a larger perturbation in the future (7-9,13).

The LoC approach proposes that many interacting regulatory processes, operating over multiple time scales, give rise to complex physiological output and allow the individual to flexibly adapt to internal and external perturbations (4,6). Age- and disease-related declines in these interactions, and the corresponding loss of physiological complexity, are associated with decreased adaptation to physiologic stress and subsequent functional decline. Thus, quantifying the complexity of physiologic systems may provide hints about the individual’s ability to adapt to future stressors. Non-linear methods like multiscale entropy (MSE), and its multivariate extension (MVMSE), can be used to quantify the complexity embedded in one or more physiological signals (14-16). Overall, lower complexity in physiological time series is generally associated with poorer functioning and lower resilience of the system under study (17,18).

Importantly, while related in their purpose, these two paradigms make use of different underlying assumptions and are hypothesized to be theoretically complementary in their potential for ascertaining resilience. CSD assumes that the small magnitude and temporal independence of fluctuations around a homeostatic equilibrium in response to perturbation indicate resilience in terms of stability, whereas LoC assumes that more complex fluctuation patterns across multiple temporal scales at rest reflect resilience in terms of adaptability (4,7,8) (see Figure 1). It is currently unknown whether, individually or combined, these indicators can assess the potential resilience of older adults.

**Figure 1:**
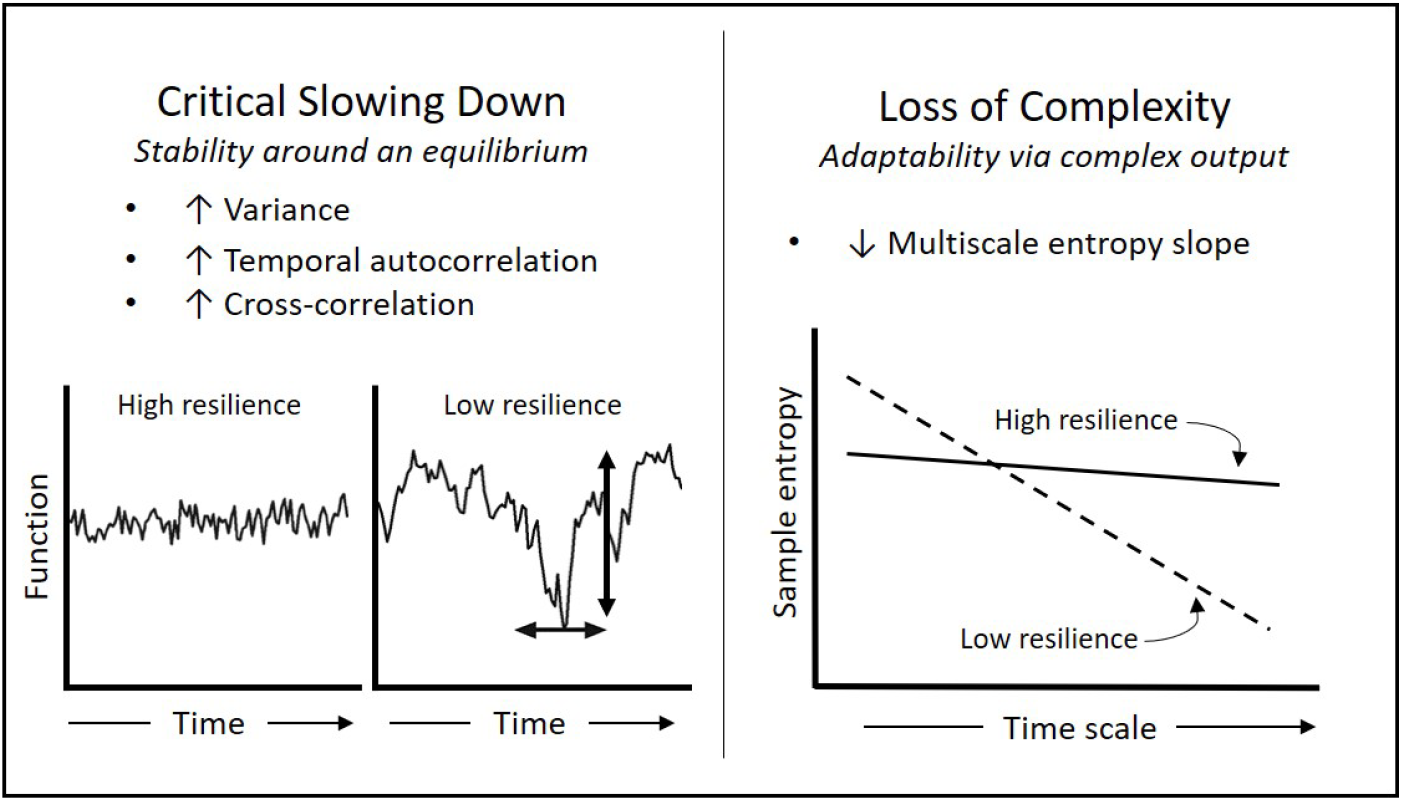
Complex systems paradigms and associated indicators quantifying resilience in physiological time series data from older adults. Critical Slowing Down (8) reflects resilience in terms of stability around an equilibrium. Indicators of low resilience from this paradigm include increased variance (vertical arrow), temporal autocorrelation (horizontal arrow) and cross-correlation between sub-systems (not shown). Loss of Complexity (4) reflects low resilience in terms of diminished adaptability of complex physiological output. Complexity can be quantified using multiscale entropy, whereby more negative slopes indicate less complexity and low resilience.

As a first step towards evaluating how dynamical resilience indicators derived from these two paradigms can reliably and validly be used in the care of acutely ill older persons, we extracted putative indicators of both paradigms using continuous heart rate and physical activity time series data collected from geriatric inpatients. Heart rate and physical activity were chosen as commonly used, dynamic variables that are relatively easy to obtain in the daily routine of patient care. To explore their construct validity, we examined the degree to which the extracted resilience indicators coincided with three measures of health functioning known to reflect greater vulnerability to stressors (i.e. low resilience) at hospital admission: frailty, multimorbidity and Activities of Daily Living (ADL). For reliability, we examined the test-retest reliability comparing LoC and CSD indicators derived from the first and second 24 hours.

## Methods

### Participants

Participants were included from the Wellbeing and Resilience Study at Radboud University Medical Center (11). Eligible patients aged 65 years or over admitted to the geriatric ward were consecutively enrolled between March and October 2017. The medical ethics committee CMO Radboudumc approved the study (ID: 2017-3225) and all participants gave written informed consent.

### Measures of health functioning

Pre-admission frailty was measured retrospectively at admission with The Older Persons and Informal Caregivers Survey – Minimal Dataset (TOPICS-MDS) 45-item frailty index (range 0-1, lower is better) (19,20). Multimorbidity was measured as the number of diseases out of a possible nineteen physician-diagnosed chronic conditions: sixteen from the TOPICS-MDS questionnaire, supplemented by information on Parkinson’s disease, aortic stenosis and cardiac rhythm disorders. The TOPICS-MDS questionnaire also results in a validated Activities of Daily Living (ADL) score (range 0-6, higher is better) (20). This score included 6 items: the ability to independently walk outside for 5 minutes, take a shower/bath, dress oneself, stand up from a chair, walk up a flight of 15 stairs, and take medication.

### Continuous monitoring of heart rate and physical activity

A chest-worn sensor, the HealthPatch MD (VitalConnect, San Jose, California, USA), was used to simultaneously collect continuous heart rate and physical activity data starting immediately after the patient was admitted to the geriatric ward. The sensor reliably sampled patients’ electrocardiogram (ECG) at 125 Hz and a triaxial accelerometer at 31.25 Hz (21). More details can be found in the Supplemental Materials.

### Heart rate and physical activity data pre-processing

Pre-processing of ECG and accelerometer data during the first 24 hours after hospital admission was carried out in MATLAB (R2014b, Mathworks, Natick, MA); details are available in the eMethods. To calculate the multivariate indicators (i.e. cross-correlation and MVMSE), the pre-processed time-series of heart rate was interpolated and re-sampled using the time stamps of the pre-processed physical activity time series. This resulted in two time-matched physical activity and heart rate signals – both sampled once every 4 seconds (0.25 Hz). Because these indicators would ideally be able to provide actionable information within the first 12 hours or less, the resilience indicators were extracted from the first 10,000 data points immediately after admission; this translated to approximately 11 hours of recording time (median[IQR]=11.1[.45] for heart rate; 11.1[.33] for physical activity). This fixed number of data points also facilitated comparison of (MV)MSE estimates between patients. As separate control variables, patients’ average heart rate (in beats per minute [bpm]) and physical activity (in activity counts per hour) were also calculated for the entire 24-hour period.

### Critical slowing down indicators calculation

Variance of heart rate and physical activity was calculated as the average squared standard deviation of the pre-processed time series from its detrended mean. Temporal autocorrelation (TAC) was calculated by correlating each time series with a time-lagged version of itself. For physical activity, the autocorrelation was calculated using a lag of 4 seconds (1 epoch). For heart rate, a lag of 1 minute was used to better capture the intrinsic dynamics of the heart. The cross-correlation was the bivariate correlation between the pre-processed heart rate and physical activity time series.

### Multiscale entropy and scaling regions

MSE was calculated using the methods described by Costa, Goldberger and Peng (15,16). Briefly, the sample entropy, a measure of information content, is calculated for progressively larger times scales within the time series. The MSE slope is the change in sample entropy values as a function of time scale and reflects the ‘meaningful structural richness’ or complexity of the time series. More negative slopes reflect less complexity, whereas more positive slopes indicate greater complexity (Figure 1). MVMSE is an extension of the MSE method to multivariate time series data (14). It reflects the joint complexity of heart rate and physical activity over time.

It is possible for time series to display distinct trends of information content across different time scales. This gives rise to so-called scaling regions of the MSE slope that may correspond to regulatory mechanisms acting within specific time scales (22). Based on visual inspection, univariate heart rate MSE and MVMSE showed two scaling regions: from scale 1-4 (4-16 seconds) and from scale 4-10 (16-40 seconds; see eFigure 1). We therefore estimated separate slopes for these two scaling regions (scaling region 1 and 2, respectively). See eMethods for more extensive methods description.

### Statistical analyses

Statistical analyses were performed in IBM SPSS version 25 (IBM Corp., Chicago, IL). To explore the unique or overlapping nature of information contained in the putative resilience indicators, bivariate correlations were carried out between the indicators of CSD and (MV)MSE slopes. Next, each of the indicators were entered as separate predictors of each of the three measures of health functioning in linear regression analyses. Each regression model was adjusted for age and sex (Model 1). To investigate whether the indicators were associated with functioning independent of average heart rate or physical activity, a second model (Model 2) was included that additionally adjusted for the corresponding 24-hour average of heart rate, physical activity or both, depending on the system of the resilience indicator being tested. As these analyses were aimed at hypothesis-driven questions of an exploratory nature, no adjustments for multiple comparisons were made and p-values should be interpreted with caution.

Three relevant supplementary analyses were carried out: relative weights analysis, a test of repeatability, and a comparison of within-person changes in indicators across paradigms. First, to assess the relative contribution of each CSD and LoC resilience indicator to explaining the variance in the measures of health functioning, relative weights analysis was conducted (23). Relative weights analysis partitions the total explained variance (R^2^) among the multiple predictors to quantify the unique contribution of each predictor in the model (24). Second, as the indicators may be prone to measurement error, we assessed within-subject test-retest reliability by repeating the main regression analyses using indicators extracted from the first 24-hour period with the indicators extracted from the second 24-hour period post-admission (24-48 hours) as independent variables. Third, if the CSD and LoC indicators indeed reflect related, but distinct information about resilience, their changes over time within-persons would show low correlations. The extraction of indicators from the second 24-hour period in the repeatability analysis also allowed for examination of whether within-person changes in indicators from CSD and LoC were correlated with one another (See eMethods for more details).

## Results

### Participant characteristics

Table 1 shows the characteristics of the 121 patients in this study. The participants’ mean ± SD age was 84.2 ± 6.3 years and 60% were female. Prior to hospital admission, 73% were living independently and 29% had a diagnosis of dementia. The average frailty index score was 0.38 ± 0.14. Participants had a mean of 4.4 ± 2.0 chronic medical conditions (multimorbidity) and an average ADL function score of 2.7 ± 2.2. For a summary of reasons for admission and most common co-morbidities, see eTable 1 and eTable 2 in the Supplemental Materials.

**Table 1:** Pre-admission characteristics of geriatric patients (n=121)

| | Mean ± SD / n (%) |
| --- | --- |
| Age (years) | 84.2 ± 6.3 |
| Females | 73 (60%) |
| Body mass index (kg/m<sup>2</sup>) | 25.2 ± 5.3 |
| Education level |  |
| Low | 60 (56%) |
| Middle | 27 (25%) |
| High | 23 (21%) |
| Living situation |  |
| Independent, alone | 52 (43%) |
| Independent, with others | 39 (32%) |
| Residential care | 18 (12%) |
| Nursing home | 12 (10%) |
| Admitted from |  |
| Emergency room | 107 (88%) |
| Elective | 9 (7%) |
| Other hospital unit | 5 (4%) |
| Dementia | 35 (29%) |
| ***Health functioning measures*** |  |
| Multimorbidity (0-19) | 4.4 ± 2.0 |
| Frailty index (0-1) | 0.38 ± 0.14 |
| ADL function (0-6) | 2.7 ± 2.2 |

Note. ADL = Activities of daily living.

**Table 2:** Resilience indicators based on two different paradigms extracted from patients’ heart rate and physical activity time series data.

|  |  | Mean | SD | N |
| --- | --- | --- | --- | --- |
| *24-hour average* |  |  |  |  |
| HR | Mean (bpm) | 83.85 | 16.75 | 112 |
| PA | Mean (counts/hour) | 625.75 | 235.87 | 119 |
| *Critical slowing down* |  |  |  |  |
| HR | Variance | 35.22 | 45.96 | 112 |
|  | TAC (1 minute) | 0.65 | 0.66 | 117 |
| PA | Variance | 0.72 | 0.17 | 102 |
|  | TAC (4 seconds) | 0.72 | 0.08 | 112 |
| HR + PA | Cross-correlation | 0.36 | 0.16 | 112 |
| *Loss of Complexity* |  |  |  |  |
| HR | Slope (scale 1-4) | 0.24 | 0.10 | 112 |
|  | Slope (scale 4-10) | 0.01 | 0.02 | 112 |
| PA | Slope (total) | -0.01 | 0.02 | 112 |
| HR + PA | Slope (scale 1-4) | 0.04 | 0.02 | 112 |
|  | Slope (scale 4-10) | 0.00 | 0.01 | 112 |

Note. HR = heart rate; PA = physical activity; HR + PA = multivariate indicator using heart rate and physical activity; TAC = Temporal autocorrelation; slope = multiscale entropy slope. More negative slopes indicate lower complexity.

### Resilience indicators

Table 2 shows the mean and standard deviation of the resilience indicators from participants’ heart rate and physical activity time series data. Of note, the MSE slope for physical activity was on average negative, but close to zero, mean ± SD = –0.01 ± 0.02 (see eFigure 1). The average MSE slope for heart rate over scaling region 1 (4-16 seconds) was positive, 0.24 ± 0.10, whereas the slope of scaling region 2 (16-40 seconds) was close to zero, 0.01 ± 0.02. Recall that more negative slopes are indicative of lower complexity, and presumably lower resilience, of the sub-system. The MVMSE displayed a similar scaling behavior as the heart rate and was also described by the same two scaling regions: a slightly positive MVMSE slope over scaling region 1, 0.04 ± 0.02 and a flat MVMSE slope for scaling region 2, 0.00 ± 0.01.

### Between-participant correlations among CSD & LoC indicators

Table 3 shows the between-participant correlations among the resilience indicators. Correlations were mostly weak-to-moderate. Comparing the two paradigms’ resilience indicators (3-7 vs. 8-12 in Table 3), 22 out of 25 correlations were below ρ=0.50 with significant associations ranging in absolute value from ρ=.197 to ρ=.472 (all p<.05). The strongest correlations between paradigms were observed for heart rate TAC (1-minute lag) and heart rate (MV)MSE slope over scaling region 1 (MSE: ρ=–.769, p<.01; MVMSE: ρ=– .694, p<.01). Physical activity variance was also moderately associated with physical activity MSE slope (ρ=.665, p=<.01).

**Table 3:**
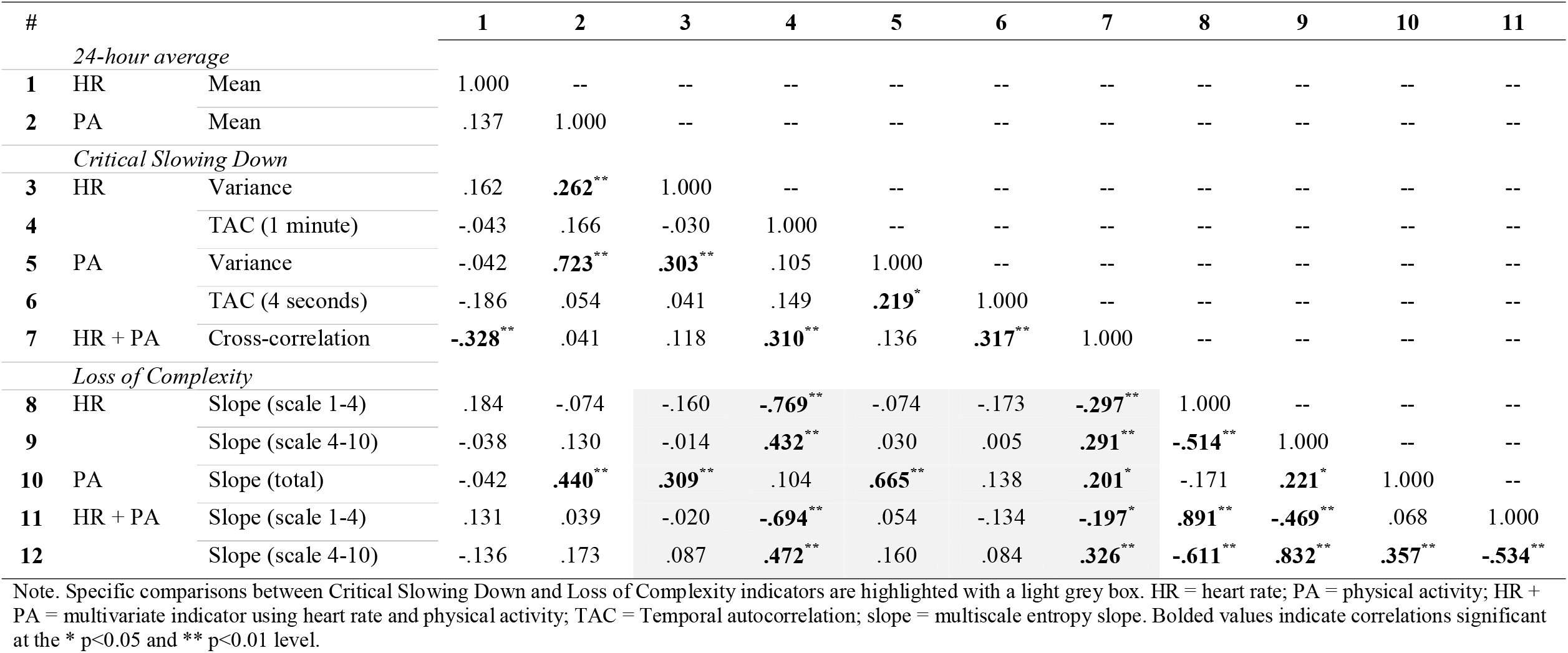
Between-participant Spearman correlation coefficients for critical slowing down and multiscale entropy indicators.

### Relationship between CSD & LoC indicators and health functioning

Table 4 shows the associations of CSD and LoC indicators with health functioning measures at admission. Higher mean 24-hour physical activity was significantly associated with lower frailty (β = –0.27, p<.010) and better ADL function (β = 0.29, p<.010). Average 24-hour heart rate was unrelated to the health functioning measures.

**Table 4:** Association between critical slowing down and multiscale entropy indicators and measures of health functioning.

|  |  | Multimorbidity |  | Frailty |  | ADL |  |
| --- | --- | --- | --- | --- | --- | --- | --- |
|  |  | Model 1 | Model 2 | Model 1 | Model 2 | Model 1 | Model 2 |
| *24-hour average* |  |  |  |  |  |  |  |
| HR | Mean | 0.12 (.220) | -- | 0.02 (.805) | -- | 0.08 (.445) | -- |
| PA | Mean | 0.01 (.911) | -- | **-0.27 (.006)\*\*** | -- | **0.29 (.004)\*\*** | -- |
| *Critical Slowing Down* |  |  |  |  |  |  |  |
| HR | Variance | 0.10 (.315) | 0.08 (.412) | -0.17 (.099) | -0.17 (.088) | 0.13 (.185) | 0.13 (.225) |
|  | TAC (1 minute) | -0.08 (.430) | -0.07 (.489) | -0.04 (.672) | -0.04 (.678) | 0.07 (.493) | 0.09 (.408) |
| PA | Variance | -0.09 (.347) | -0.27 (.068) | **-0.37 (<.001)\*\*\*** | -0.44 (.006) | **0.38 (<.001)\*\*\*** | **0.42 (.008)\*\*** |
|  | TAC (4 seconds) | -0.02 (.853) | -0.02 (.845) | -0.15 (.134) | -0.13 (.191) | **0.20 (.050)\*** | 0.17 (.076) |
| HR + PA | Cross-correlation | -0.06 (.508) | -0.02 (.860) | -0.07 (.460) | -0.07 (.520) | 0.01 (.907) | 0.05 (.663) |
| *Loss of Complexity* |  |  |  |  |  |  |  |
| HR | Slope (scale 1-4) | 0.15 (.111) | 0.13 (.175) | 0.14 (.152) | 0.15 (.157) | -0.09 (.395) | -0.11 (.299) |
|  | Slope (scale 4-10) | **-0.24 (.010)\*\*** | **-0.23 (.016)\*** | -0.10 (.317) | -0.10 (.330) | -0.03 (.752) | -0.02 (.843) |
| PA | Slope (total) | -0.04 (.691) | -0.05 (.626) | **-0.28 (.004)\*\*** | -0.23 (.019) | **0.23 (.022)\*** | 0.17 (.080) |
| HR + PA | Slope (scale 1-4) | 0.14 (.137) | 0.13 (.201) | 0.12 (.245) | 0.11 (.274) | -0.11 (.290) | -0.12 (.244) |
|  | Slope (scale 4-10) | **-0.24 (.011)\*** | **-0.23 (.019)\*** | -0.17 (.081) | -0.16 (.109) | 0.07 (.480) | 0.08 (.456) |

Note: Values are standardized beta coefficient (p-value); Model 1 adjusted for age and sex; Model 2 = Model 1 additionally adjusted for mean heart rate, mean physical activity or both. ADL = Activities of daily living; HR = heart rate; PA = physical activity; HR + PA = multivariate indicator using heart rate and physical activity; TAC = Temporal autocorrelation; slope = multiscale entropy slope. Bolded coefficients significant at the \* p<0.05, \*\* p<0.01 and \*\*\* p<0.001 level.

More positive MSE slopes, indicative of higher complexity, for physical activity were associated with lower frailty index scores (β = –0.28, p=.004) and better ADL function (β = 0.23, p=.022). The latter association with ADL function was reduced to a trend after adjustment for mean 24-hour physical activity. More positive heart rate MSE slopes were associated with lower multimorbidity for scaling region 2 (16-40 seconds) only (β = –0.24, p=.010). The pattern for the MVMSE slopes was nearly identical to that found for the univariate heart rate MSE slope.

Opposite to the assumptions under CSD, greater variance in physical activity was associated with lower frailty index scores (β = –0.37, p<.001) and better ADL function (β = 0.38, p<.001). Increased temporal autocorrelation in physical activity was likewise associated with better ADL function (β = 0.20, p=.050). This latter association with ADL function was attenuated after adjustment for mean 24-hour physical activity. The remaining associations were non-significant (all p>.05).

### Relative weights, repeatability, and associations of within-person change

The results of the relative weights analysis and repeatability analysis are available in the Supplemental Materials. Briefly, relative weights analyses revealed that three indicators derived from physical activity – the 24-hour average, variance and MSE slope – were the top 3 contributors to the explained variance in frailty index and ADL function. Specifically, physical activity variance accounted for a significant proportion of the total explained variance (R^2^) in frailty (35%) and ADL function (38%). The reliability analyses showed that most of the indicators were relatively stable from the first to the second day of hospitalization. Finally, the associations of within-person changes between CSD and LoC indicators revealed nearly all weak (i.e. below ρ = |0.20|) and non-significant (p > 0.50) associations, providing further support that these paradigms reflect related, but distinct aspects of resilience (see eResults and eTables 3-6).

## Discussion

The general lack of strong correlations between CSD and LoC indicators suggested that they represent largely unique resilience features embedded within the time series. Greater physical activity variance and complexity were significantly associated with lower frailty index scores and greater ADL function, while greater heart rate complexity was associated with lower multimorbidity. In this group of acutely ill geriatric inpatients, there were no associations in the hypothesized direction between resilience indicators based on the CSD paradigm and health functioning. In fact, contrary to the theory of CSD, increased physical activity variance was associated with lower instead of higher frailty and disability. Here, we will scrutinize several factors related to how we used the two paradigms that – in retrospect – may offer plausible explanations for the pattern of associations. Importantly, we provide a number of lessons learned for other researchers interested in applying these paradigms to the resilience of ill older persons.

### Homeostatic roles of regulated and effector variables

When designing this study, we did not consider one aspect that we later found is crucial for empirically evaluating indicators from the CSD and LoC paradigms. That is the distinction between the homeostatic roles of regulated variables, such as blood pressure, which need to be maintained within tight ranges, and effector variables, such as heart rate, which flexibly adapt to perturbations to help maintain the regulated variables within their physiological bounds (25). For regulated variables, high variability may reflect loss of resilience, whereas for effector variables, high variability may reflect greater resilience. Considering these homeostatic roles, it is proposed that the CSD indicators primarily reflect loss of stability in regulated variables fluctuating around an equilibrium, whereas LoC indicators best reflect loss of adaptability in effector variables (25,26).

We included physical activity and heart rate as commonly used, easily obtainable time series that also reflect aspects of homeostasis and systemic resilience, given their crucial involvement in the maintenance of health and response to illness. There is certainly a well-established link between average physical activity levels and health functioning, including health in mental and social domains (27-30). Similarly, indicators derived from cardiovascular dynamics can discriminate patients with various heart conditions from healthy controls and help understand the breakdown in heart regulation with disease (31,32). While at longer time scales of days to weeks, heart rate and physical activity conceivably fluctuate around an equilibrium, this is less obvious for the shorter time scales of seconds to minutes considered in this study. At this resolution, neither physical activity nor heart rate can be reasonably considered to behave as regulated variables. Applying the CSD indicators to these variables may have thus been inappropriate based on their expected behavior and the underlying assumptions of the paradigm.

Taken together, these findings suggest that it is important to match the underlying assumptions of each paradigm to the homeostatic role of the variable one uses to extract potential indicators of resilience in geriatric patients. Other sub-systems than the ones studied here, such as the balance system or blood pressure are shown to be more appropriate for applying CSD indicators (33). In addition to measures of these sub-systems at rest, provocative tests like the orthostatic (sit-to-stand) challenge can further probe resilience and provide deeper insight, if not contraindicated for the patient.

### Intuitive associations with physical activity

The association of activity-derived MSE slopes with health functioning is consistent with previous findings linking physical activity complexity with advanced age and mortality in older adults using other complexity methods (34). However, the positive association between physical activity variability and health functioning may be expected intuitively given that more physical activity in hospitalized adults is typically a good sign (except when due to restlessness or acute confusion). For example, serious ailments requiring hospitalization are often associated with reduced mobility (35,36). Patients with higher average levels of physical activity, despite their condition, would thus be expected to display more (not less) physical activity variance and have greater health functioning. This idea is supported by the sizable positive correlation between mean 24-hour activity and variance in this study (ρ=.723, p<.001). This opposing finding is likely unrelated to presence or absence of CSD and tells more about the function of the physical activity than the validity of the paradigm. Future tests of these paradigms should consider whether the hypothesized directions of associations are in line with the nature of the sub-system measured.

### Noise inherent in empirical time series data

Real data are often very noisy, certainly free-living time series of persons with many deficits and morbidities. For example, approximately one third (n=35, 31%) of the current study population reported a cardiac rhythm disorder, including atrial fibrillation. These conditions may have impacted the relationships observed with indicators derived from the heart rate time series data. However, despite these conditions, increased heart rate complexity was still associated with lower multimorbidity, even after additional adjustment. Although typically done in comparable studies (37,38), excluding these patients would have strongly biased the results, as well as hindered generalizability. Thus, this could also be considered a strength of this study. Considering the types of noise expected, its potential sources and ways to prevent it, if possible, will undoubtedly improve the insights garnered from the use of these indicators in future research. It is also possible, for example, to apply the indicators to time series data obtained from patients under more controlled conditions, including standardized fatigability tests like grip work assessment (39).

### Unknown trajectories of geriatric inpatients

Patients admitted to the hospital, by definition, have experienced a stressor too great to resist. Except for planned stressors (e.g., elective surgery), patient monitoring is not synchronized with the stressor but often starts from an arbitrary moment within the time course of the patient’s condition. Therefore, the decrement from pre-stressor functioning and the phase of the recovery process that the patient is currently in are largely unknown. The considerable heterogeneity in the patients’ diagnoses and in the corresponding time course of their resolution means that this study likely assessed snapshots of individuals at different phases of the disease process. While comparing geriatric patients’ relative resilience may help identify those most in need of care, it cannot reveal whether an individual patient’s condition is worsening or improving. An alternative approach is to perform regular measurements in a large, longitudinal cohort and wait for stressors to occur, paying close attention to within-rather than between-person changes in indicators. Particularly, studies involving multiple repeated measures over a prolonged period that adaptively include higher-frequency measurements around a stressor (e.g., ‘measurement-burst’ study designs) are scarce but valuable (40). Examining resilience indicators obtained from such study designs may better discern the within-person dynamics in response to various health stressors.

### Unknown stable states of geriatric inpatients

From the perspective of the CSD paradigm, it is not uncommon for a strong perturbation to force an adaptive system functioning in one state (e.g., healthy) to shift into an another stable, albeit less desirable state (e.g., disease). This new, alternative state may be quite resilient, as can be seen in reinforcing feedback loops of pathophysiological or psychiatric symptom networks (7,13). Importantly, the CSD resilience indicators reflect the resilience of the state that the system is in and may therefore reflect a high resilience for patients that are in a diseased state. Thus, another consequence of taking only snapshots of patients at hospitalization is that we cannot differentiate patients that are settling back into their ‘normal’, healthy state from those that – due to the health stressor – have moved into an alternative, equally stable ‘disease’ state. Consideration of other clinical characteristics should help here.

### Strengths and limitations

There are still other features of the current study that should be highlighted. Importantly, with the recent surge in time-intensive data from wearable sensors, there is a need for techniques that reliably extract actionable indicators that can improve clinical decision-making. A major strength of this study is that we took a theory-based approach that translates features embedded in physiological time series data into potential indicators of resilience in the face of a health stressor. While attempting to put the indicators to the test of empirical validation, we discovered potential pitfalls and tips for their proper application not typically discussed in the current literature.

Beyond the lessons learned above, another limitation is the cross-sectional study design, which is especially relevant for the measurement of resilience as an inherently dynamic process. As a first step towards evaluating the construct validity of these indicators in geriatric inpatients, resilience was proxied here by static measures reflecting increased vulnerability to health stressors. However, longitudinal study designs will facilitate comparison of these indicators to patients’ resilience trajectories (e.g., (41)) and provide insight into their ability to predict the dynamic actualization of resilience in older adults.

## Conclusion

The current study provided evidence that, in a heterogeneous sample of geriatric inpatients, indicators from Critical Slowing Down and Loss of Complexity capture largely distinct underlying features from physiological time series data. Our findings stress that different or even opposing patterns of associations may arise when applying different indicators to regulated or effector variables and further emphasizes the importance of matching the paradigms’ underlying assumptions. These results warrant the further development of an overarching framework that will guide researchers and allow medicine to leverage the most appropriate tools to gain insight into resilience using time series data from both regulated and effector variable types. Future studies should include both regulated and effector variable types and directly investigate how well these indicators predict resilience, operationalized as the dynamic trajectory of geriatric patients, as well as their added value above and beyond traditional non-dynamic measures of health functioning.

## Data Availability

Data can be made available upon request.

## Funding

None.

### Acknowledgements

The authors thank J. Zhou for supplying statistical coding.

