## Supplemental Materials for "Dynamical Indicators of Resilience from Physiological Time Series in Geriatric Inpatients: Lessons Learned"

**Contents**

- eMethods
- eResults
- eFigure 1
- eFigure 2
- eTable 1
- eTable 2
- eTable 3
- eTable 4
- eTable 5
- eTable 6

**eMethods**

*Continuous monitoring of heart rate and physical activity.* The chest-worn sensor, the HealthPatch MD (VitalConnect, San Jose, California, USA), contains electrodes sampling an electrocardiogram (ECG) at 125 Hz and a triaxial accelerometer sampling at 31.25 Hz. The sensor reliably measures heart rate by calculating RR-intervals with an algorithm based on automated detection of QRS-complexes from the ECG waveforms (1). The sensor can continuously monitor the patient for 3-5 days, depending on the battery life. The sensor module recorded the incoming signals and transmitted the data via Bluetooth to an iPod (Apple, Cupertino, California, USA) for temporary storage before being transferred to the central data repository.

*Heart rate and accelerometer data pre-processing*. Pre-processing of ECG and accelerometer data was carried out in MATLAB (R2014b, Mathworks). Inter-beat intervals as detected by the sensor system were converted to heart rate in beats per minute. To ensure generalizability of the study results, the 35 participants who reported a cardiac rhythm disorder (31% of the study sample) were not excluded from heart rate analyses. A reliable minute-to-minute heart rate signal was established by smoothing the raw heart rate (beats per minute) signal by locally weighted scatterplot smoothing (LOESS) with a span of 60 seconds (‘rloess’ function in MATLAB R2014b). To facilitate calculation of Critical Slowing Down indicators, heart rate data for the first 24 hours were additionally detrended using a span of 12 hours.

Accelerations were stored in gravity (g) units (1 g = 9.81 m/s²). The mean vector magnitudes – calculated as the Euclidean norm (square root of the sum squared raw acceleration of all three mutually perpendicular axes) – were filtered with a fourth order, high-pass filter with cut-off frequency of 0.1 Hz to remove very slow sensor drifts (2). This resulted in a time series centered around zero. Next, the absolute value was taken to obtain all positive vector magnitudes. Finally, the time series was converted into counts per 4-second epoch by summing all vector magnitudes within non-overlapping consecutive 4-second windows (2).

Those missing data for more than 9 hours of the first 24-hour period were excluded. Missing data for the remaining time series were handled differently depending on the indicator being calculated. For variance, missing data was excluded before calculation. For temporal autocorrelation, gaps in the signal larger than 60 seconds were considered too large for a reliable measure. In this case, temporal autocorrelation was calculated for each of the individual segments longer than 60 consecutive data points and the total temporal autocorrelation of the signal was a sum weighted by segment length. For the (multivariate) multiscale entropy, missing data points were first removed, and the resultant sample entropy value was weighted according to the amount of missing data.

*Multiscale entropy and scaling regions*. Briefly, MSE comprises two steps: a course-graining procedure and subsequent sample entropy calculation on the course-grained time series. For a given time series, multiple coarse-grained time series are constructed by averaging the data points within non-overlapping windows of increasing length corresponding to the time scale (called the scale factor). For each of these course-grained time series, the sample entropy – a measure that quantifies the predictability of the time series – is calculated and plotted as a function of the scale factor. Sample entropy estimates the conditional probability that two sequences of m consecutive data points, which are similar to each other within a given tolerance (r * SD) will remain similar when considering *m*+1 consecutive points. In the current study, these two user-defined parameters were set as m=2, and r=0.15. The MATLAB sample entropy implementation was obtained from PhysioNet (3,4).

The current study examined MSE over 10 scale factors. Scale 1 corresponds to the original data, which in the current study is 4 seconds. Scale 2 is 8 seconds, and so forth, up to scale 10 which is 40 seconds. The slope of sample entropy values as a function of scale factor, therefore, quantifies the complexity of the time series across multiple time scales. MVMSE is the multivariate extension of MSE. The same principle applies as in the univariate version, but instead uses information from multiple variables to calculate the entropy while accounting for both within- and cross-channel dependencies of the multivariate signals. In other words, it reflects the joint complexity of heart rate and physical activity over time.

To assess whether the frailty level itself influenced the MSE curve, stratified MSE curves for heart rate and physical activity were plotted using the mean frailty score (0.37) as a cut-off for a high- and low-frailty groups (see eFigure 2). This plot revealed that there were no distinguishable differences between those above and below the mean frailty score for heart rate or physical activity.

*Relative weights analysis*. Relative weight analysis is a useful supplement to multiple regression analysis that helps address multicollinearity issues when independent variables are correlated with one another and partitions the total explained variance (R^2^) among multiple predictors to quantify the unique contribution of each predictor in the model (5,6). The resulting percentages, therefore, represent the relative proportion of the total R^2^ accounted for by each individual resilience indicator.

*Test-retest reliability*. To assess the within-subject test-retest reliability, the main regression analyses were repeated using indicators extracted from the second 24-hour period post-admission (24-48 hours) as independent variables. First, the identical procedures were used to extract the indicators from data from the second 24-hour period (day 2) of hospitalization, which were then compared to indicators from the first 24-hour period (day 1) by t-tests and correlations analyses. Second, the indicators from day 2 were entered into identical linear regression models as in the main analyses.

*Associations of within-person change*. If the CSD and LoC indicators reflect related, but distinct information about resilience, their changes over time within-persons would show negligible correlations. Using the values for the indicators extracted across the two consecutive 24-hour periods, a comparison of the within-person changes in indicators was additionally made. First, a difference score was calculated between day 1 and day 2 indicators, representing within-person change. Then, the difference scores for each Critical Slowing Down (CSD) indicator was correlated with each Loss of Complexity (LoC) indicator.

**eResults**

*Reasons for admission and common comorbidities*. eTable 1 summarizes the primary diagnoses of the n=121 participants of the study. The most prevalent reason for admission was infection (29%) followed by functional decline or a fall (16%). The next prevalent reasons were hip fracture/surgery (13%) and cardiac issues (13%). eTable 2 summarizes the most common co-morbidities. The most commonly experienced was incontinence (57%), followed by sensory problems (hearing and vision; 43% and 38%, respectively).

*Relative weights analysis*. eTable 3 shows the results of the relative weights analysis. Using each measure of health functioning as a separate dependent variable, the same variables that were included in Model 2 (Table 4) of the main analyses (i.e. age, gender, 24-hour average heart rate and physical activity, as well as all CSD and (MV)MSE indicators) were entered as independent variables in a single model. Each predictor’s respective contribution to the total R^2^ was then estimated. These analyses revealed that three indicators derived from physical activity – the 24-hour average, variance and MSE slope – were the top 3 contributors to the explained variance in the frailty index and ADL function. Specifically, physical activity variance accounted for a significant proportion of the explained variance in frailty (35%) and ADL function (38%). No other resilience indicators were significant contributors to the explained variance in any one health functioning measure.

*Repeatability between first- & second-day estimates*. eTable 4 shows the differences and correlations between day 1 and day 2 resilience indicators. While most of the indicators appeared to be stable, the average variance and average temporal autocorrelation of physical activity was significantly higher on day 2 compared to day 1 of hospitalization. Except for heart rate variance (ρ=0.272, p=.006), the correlations of the indicators with their next-day values ranged from ρ=0.608 to ρ=0.871 (all p<.001).

eTable 5 shows the comparison of the fully adjusted regression model (Model 2) using day 1 and day 2 indicators. The pattern of results, in terms of direction and magnitude, was mostly consistent across both days. The day 1 relationships, including the associations of variance in physical activity with frailty index and ADL function, as well as the association of physical activity MSE slope with frailty remained for day 2. Relationships that were previously non-significant between physical activity variance and multimorbidity, heart rate variance and frailty index as well as physical activity MSE slope and ADL strengthened from a trend to significantly associated on day 2. Conversely, the relationships of multimorbidity with scaling regions 2 for heart rate MSE and MVMSE on day 1 were no longer significant on day 2. Overall, these results suggest that the resilience indicators are stable over time, have relatively consistent associations with health functioning or may become associated within the first 48 hours of hospitalization.

eTable 6 shows the correlations comparing within-person changes in CSD indicators with changes in LoC indicators from day 1 to day 2. These analyses showed that nearly all correlations between changes in CSD and LoC indicators were very weak (i.e. below ρ = |0.20|) and non-significant (p > 0.50). One exception was found for the heart rate-derived indicators whereby within-person decreases in MSE slope (for scaling region 1 only) was associated with increases in variance and temporal autocorrelation (variance: ρ = – 0.643; autocorrelation: ρ = – 0.662; both p < 0.001). A similar pattern was found for the MVMSE, but once again, this is likely related to the dominance of the heart rate signal in the MVMSE indicator.

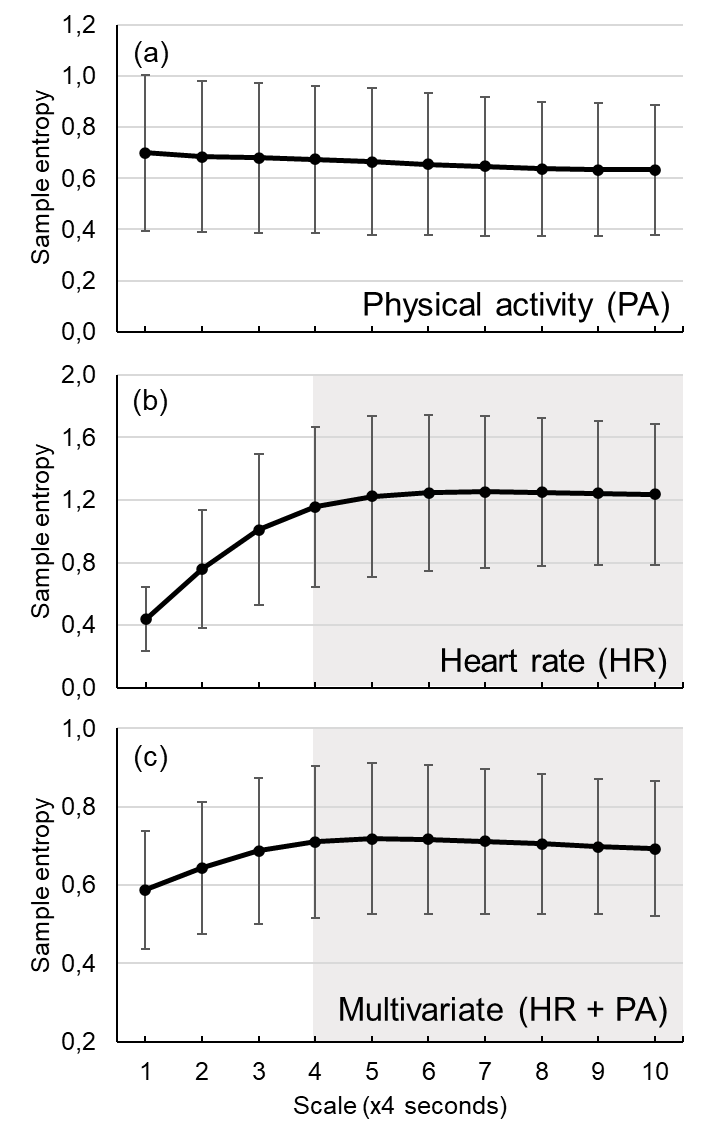

**eFigure 1**: Average multiscale entropy curves for (a) physical activity, (b) heart rate and (c) multivariate heart rate and physical activity. Error bars are standard deviation. Scaling region 1 and 2 for heart rate and the multivariate case are distinguished by a light grey box.

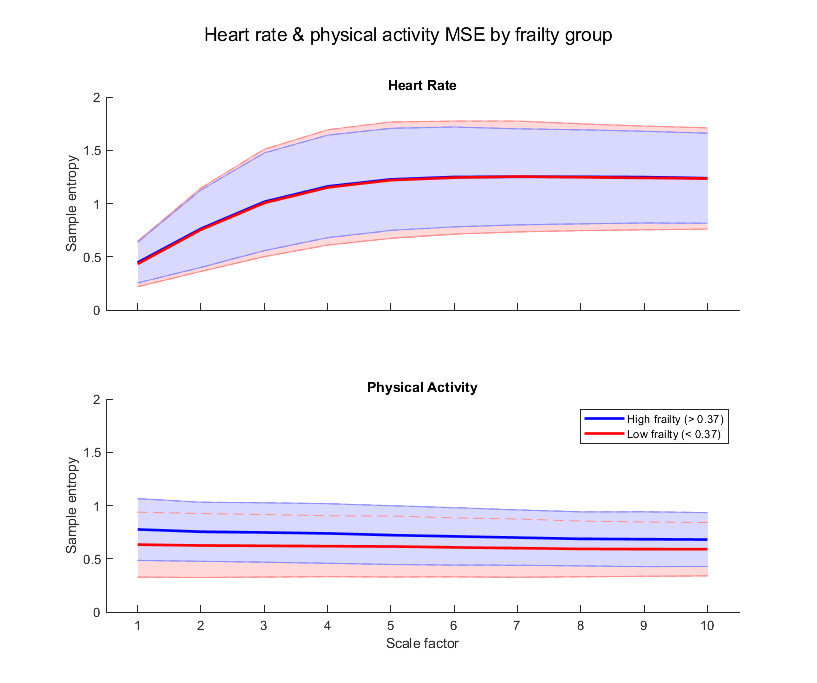

**eFigure 2**: Average multiscale entropy (MSE) curves for heart rate (top) and physical activity (bottom) stratified by frailty group. High and low frailty groups had values above and below the average, 0.37, respectively. Shaded bands are standard deviation.

| **eTable 1: Primary hospital diagnoses (n=121)** | |
| --- | --- |
|  | **n (%)** |
| Infection | 35 (29%) |
| Functional decline or fall | 19 (16%) |
| Hip fracture / surgery | 16 (13%) |
| Cardiovascular | 16 (13%) |
| Cancer | 6 (5%) |
| New dementia diagnosis | 5 (4%) |
| Dehydration | 4 (3%) |
| Pulmonary | 3 (2%) |
| Stroke | 3 (2%) |
| Other | 14 (12%) |

| **eTable 2: Most common co-morbidities** | |
| --- | --- |
|  | **n (%)** |
| Involuntary urinary loss (incontinence) | 69 (57%) |
| Hearing problems | 53 (44%) |
| Vision disorders | 46 (38%) |
| Heart failure | 41 (34%) |
| Dementia | 35 (29%) |
| Joint damage (osteoarthritis, rheumatoid wear) of hips or knees | 34 (28%) |
| Diabetes | 32 (26%) |
| Stroke, brain haemorrhage, cerebral infarction | 27 (22%) |
| Respiratory disorders (asthma, chronic bronchitis, emphysema, COPD) | 26 (21%) |
| Dizziness with falling | 25 (21%) |
| A form of cancer (malignant disease) | 20 (17%) |
| Osteoporosis (osteoporosis) | 19 (16%) |
| Hip fracture | 19 (16%) |
| Fractures other than hip | 17 (14%) |
| Prostatism due to benign prostatic hyperplasia | 14 (12%) |
| Depression | 13 (11%) |
| Anxiety / panic disorder | 10 (8%) |
| Note. COPD = chronic obstructive pulmonary disorder | |

| **eTable 3: Relative weights analysis partitioning the total explained variance in measures of health functioning among the indicators** | | | | | | | |
| --- | --- | --- | --- | --- | --- | --- | --- |
|  |  | **Multimorbidity** | | **Frailty** | | **ADL** | |
|  |  | **Explained variance^a^** | **Relative %^b^** | **Explained variance** | **Relative %** | **Explained variance** | **Relative %** |
|  | Age | 0.033 (0.003 - 0.104) | **20.32** | 0.003 (0.000 - 0.016) | 1.47 | 0.001 (0.000 - 0.001) | 0.50 |
|  | Sex | 0.002 (0.000 - 0.009) | 1.42 | 0.010 (0.001 - 0.059) | 5.33 | 0.002 (0.000 - 0.003) | 1.06 |
| *24-hour average* | |  |  |  |  |  |  |
| HR | Mean | 0.001 (0.000 - 0.000) | 0.44 | 0.001 (0.000 - 0.001) | 0.68 | 0.008 (0.001 - 0.064) | 4.43 |
| PA | Mean | 0.012 (0.002 - 0.045) | 7.30 | 0.026 (0.006 - 0.074) | **14.12** | 0.027 (0.007 - 0.077) | **14.47** |
| *Critical Slowing Down* | |  |  |  |  |  |  |
| HR | Variance | 0.011 (0.001 - 0.070) | 6.63 | 0.012 (0.001 - 0.043) | 6.65 | 0.006 (0.001 - 0.023) | 3.26 |
|  | TAC (1 minute) | 0.002 (0.000 - 0.002) | 1.24 | 0.003 (0.000 - 0.006) | 1.67 | 0.002 (0.000 - 0.001) | 0.89 |
| PA | Variance | 0.026 (0.004 - 0.076) | **15.92** | 0.064 (0.015 - 0.148) | **35.06*** | 0.070 (0.018 - 0.156) | **37.78*** |
|  | TAC (4 seconds) | 0.001 (0.000 - 0.001) | 0.41 | 0.005 (0.001 - 0.029) | 2.81 | 0.017 (0.002 - 0.086) | 9.21 |
| HR + PA | Cross-correlation | 0.001 (0.000 - 0.000) | 0.34 | 0.004 (0.000 - 0.012) | 2.12 | 0.002 (0.000 - 0.003) | 1.18 |
| *Loss of Complexity* | |  |  |  |  |  |  |
| HR | Slope (scale 1-4) | 0.006 (0.001 - 0.011) | 3.82 | 0.005 (0.001 - 0.008) | 2.54 | 0.004 (0.000 - 0.007) | 2.41 |
|  | Slope (scale 4-10) | 0.034 (0.004 - 0.092) | **20.79** | 0.003 (0.000 - 0.007) | 1.89 | 0.010 (0.001 - 0.037) | 5.35 |
| PA | Slope (total) | 0.008 (0.002 - 0.040) | 4.84 | 0.033 (0.006 - 0.095) | **18.07** | 0.021 (0.004 - 0.074) | **11.31** |
| HR + PA | Slope (scale 1-4) | 0.004 (0.001 - 0.006) | 2.51 | 0.003 (0.001 - 0.005) | 1.86 | 0.006 (0.001 - 0.014) | 3.14 |
|  | Slope (scale 4-10) | 0.023 (0.004 - 0.056) | 14.01 | 0.010 (0.001 - 0.035) | 5.74 | 0.009 (0.001 - 0.022) | 5.00 |
|  | ***Total R^2^*** | 0.162 |  | 0.183 |  | 0.186 |  |
| Note. Bolded values are the three highest contributors to the total R^2^; ADL = Activities of daily living; HR = heart rate; PA = physical activity; TAC = temporal autocorrelation; slope = multiscale entropy slope. ^a^ Variance explained in the dependent variable (95% confidence interval); ^b^ Proportion of total R^2^ accounted for by the independent variable; *Contribution to total R^2^ significantly different from zero. | | | | | | | |

| **eTable 4: Comparison between indicators derived from day 1 (0-24 hours) and day 2 (24-48 hours) time series** | | | | | | | | |
| --- | --- | --- | --- | --- | --- | --- | --- | --- |
|  |  | **N** | **Day 1** | **Day 2** | **Difference** | **p-value** | **Correlation^a^** | **p-value** |
| *24-hour average* | |  |  |  |  |  |  |  |
| HR | Mean | 102 | 84.03 (17.00) | 84.56 (16.51) | 0.52 (8.53) | .537 | 0.871 | <.001 |
| PA | Mean | 118 | 624.29 (236.33) | 641.26 (251.62) | 16.98 (179.97) | .308 | 0.730 | <.001 |
| *Critical Slowing Down* | |  |  |  |  |  |  |  |
| HR | Variance | 102 | 34.25 (46.94) | 37.3 (26.77) | 3.05 (47.3) | .517 | 0.272 | .006 |
|  | TAC (1 minute) | 102 | 0.72 (0.17) | 0.72 (0.19) | 0.00 (0.13) | .816 | 0.738 | <.001 |
| PA | Variance | 111 | 0.64 (0.66) | 0.75 (0.79) | 0.11 (0.47) | **.016** | 0.808 | <.001 |
|  | TAC (4 seconds) | 107 | 0.72 (0.08) | 0.74 (0.08) | 0.02 (0.07) | **.006** | 0.608 | <.001 |
| HR + PA | Cross-correlation | 102 | 0.35 (0.16) | 0.37 (0.16) | 0.02 (0.13) | .175 | 0.642 | <.001 |
| *Loss of Complexity* | |  |  |  |  |  |  |  |
| HR | Slope (scale 1-4) | 102 | 0.24 (0.10) | 0.23 (0.11) | -0.01 (0.06) | .290 | 0.817 | <.001 |
|  | Slope (scale 4-10) | 102 | 0.01 (0.02) | 0.01 (0.02) | 0.00 (0.02) | .167 | 0.741 | <.001 |
| PA | Slope (total) | 111 | -0.01 (0.02) | -0.01 (0.02) | 0.00 (0.01) | .571 | 0.670 | <.001 |
| HR + PA | Slope (scale 1-4) | 102 | 0.04 (0.02) | 0.04 (0.02) | 0.00 (0.02) | .268 | 0.733 | <.001 |
|  | Slope (scale 4-10) | 102 | 0.00 (0.01) | 0.00 (0.01) | 0.00 (0.01) | .166 | 0.682 | <.001 |
| Note. Values are mean (SD). HR = heart rate; PA = physical activity; TAC = temporal autocorrelation; slope = multiscale entropy slope. ^a^ Spearman correlation coefficient. | | | | | | | | |

| **eTable 5: Association of critical slowing down and multiscale entropy indicators from day1 (0-24 hours) and day 2 (24-48 hours) with measures of health functioning** | | | | | | | |
| --- | --- | --- | --- | --- | --- | --- | --- |
|  |  | **Multimorbidity** | | **Frailty** | | **ADL** | |
|  |  | **Day 1** | **Day 2** | **Day 1** | **Day 2** | **Day 1** | **Day 2** |
| *Critical Slowing Down* | |  |  |  |  |  |  |
| HR | Variance | 0.08 (.412) | -0.09 (.368) | -0.17 (.088) | **-0.21 (.048)*** | 0.13 (.225) | 0.18 (.099) |
|  | TAC (1 minute) | -0.07 (.489) | -0.12 (.236) | -0.04 (.678) | -0.10 (.347) | 0.09 (.408) | 0.12 (.284) |
| PA | Variance | -0.27 (.068) | **-0.32 (.023)*** | **-0.44 (.006)**** | **-0.43 (.002)*** | **0.42 (.008)**** | **0.32 (.025)*** |
|  | TAC (4 seconds) | -0.02 (.845) | -0.07 (.439) | -0.13 (.191) | -0.10 (.295) | 0.17 (.076) | 0.06 (.529) |
| HR + PA | Cross-correlation | -0.02 (.860) | -0.13 (.235) | -0.07 (.520) | -0.03 (.778) | 0.05 (.663) | 0.02 (.848) |
| *Loss of Complexity* | |  |  |  |  |  |  |
| HR | Slope (scale 1-4) | 0.13 (.175) | 0.14 (.158) | 0.15 (.157) | 0.14 (.203) | -0.11 (.299) | -0.13 (.213) |
|  | Slope (scale 4-10) | **-0.23 (.016)*** | -0.15 (.138) | -0.10 (.330) | 0.01 (.921) | -0.02 (.843) | -0.08 (.469) |
| PA | Slope (total) | -0.05 (.626) | -0.16 (.112) | **-0.23 (.019)*** | **-0.38 (<.001)***** | 0.17 (.080) | **0.33 (<.001)***** |
| HR + PA | Slope (scale 1-4) | 0.13 (.201) | 0.07 (.465) | 0.11 (.274) | 0.01 (.889) | -0.12 (.244) | -0.05 (.614) |
|  | Slope (scale 4-10) | **-0.23 (.019)*** | -0.05 (.653) | -0.16 (.109) | 0.02 (.878) | 0.08 (.456) | -0.05 (.610) |
| Note. Values are standardized beta coefficients (p-value). ADL = Activities of daily living; HR = heart rate; PA = physical activity; TAC = temporal autocorrelation; slope = multiscale entropy slope. Models adjusted for age and sex and mean HR, PA or both (for HR + PA models). Significant at the * p<0.05, ** p<0.01 and *** p<0.001 level. | | | | | | | |

| **eTable 6: Spearman correlations coefficients comparing within-person changes from the first to second 24-hour period between Critical Slowing Down and Loss of Complexity indicators.** | | | | | | | | |
| --- | --- | --- | --- | --- | --- | --- | --- | --- |
|  |  | HR | | | PA | HR + PA | | |
|  | | Δ Slope  (scale 1-4) | Δ Slope  (scale 4-10) | Δ Slope  (total) | | | Δ Slope  (scale 1-4) | Δ Slope  (scale 4-10) |
| HR | Δ Variance | **-.643^**^** | .160 | .100 | | | **-.335^**^** | .078 |
|  | Δ TAC (1 minute) | **-.662^**^** | .110 | .021 | | | **-.392^**^** | .140 |
| PA | Δ Variance | .006 | -.062 | .067 | | | .011 | -.022 |
|  | Δ TAC (4 seconds) | .161 | .075 | -.031 | | | .118 | -.052 |
| HR + PA | Δ Cross-correlation | -.159 | .018 | .048 | | | -.007 | -.083 |
| Note. Critical Slowing Down indicators are in the rows and Loss of Complexity indicators are in the columns. HR = heart rate; PA = physical activity; HR + PA = multivariate indicator using heart rate and physical activity; TAC = temporal autocorrelation; slope = multiscale entropy slope. Bolded values indicate correlations significant at the ** p<0.01 level. | | | | | | | | |
